# Efficacy of Convalescent Plasma Therapy compared to Fresh Frozen Plasma in Severely ill COVID-19 Patients: A Pilot Randomized Controlled Trial

**DOI:** 10.1101/2020.10.25.20219337

**Authors:** Meenu Bajpai, Suresh Kumar, Ashish Maheshwari, Karan Chhabra, Pratibha kale, Amita Gupta, Ashad Narayanan, Ekta Gupta, Nirupama Trehanpati, Chhagan Bihari, Reshu Agarwal, Kamini Gupta, Upendra kumar Gupta, Ankit Bhardwaj, Guresh Kumar, Mojahidul Islam, Ravinder Singh, Pushpa Yadav, Rakhi Maiwall, Shiv Kumar Sarin

**Affiliations:** Department of Transfusion Medicine, Institute of Liver and Biliary Sciences, New Delhi, India; Department of Medicine, Lok Nayak Jai Prakash Hospital, New Delhi, India; Department of Microbiology, Institute of Liver and Biliary Sciences, New Delhi, India; Department of Hepatology, Institute of Liver and Biliary Sciences, New Delhi, India; Department of HepatoVirology, Institute of Liver and Biliary Sciences, New Delhi, India; Department of Molecular and Cellular Medicine, Institute of Liver and Biliary Sciences, New Delhi, India; Department of Pathology, Institute of Liver and Biliary Sciences, New Delhi, India; Masters Clinical Research, Department of Clinical Research, Institute of Liver and Biliary Sciences, New Delhi, India; Department of Research, Institute of Liver and Biliary Sciences, New Delhi, India

**Keywords:** COVID-19, Convalescent Plasma, Donor Plasmapheresis, ARDS, Immunoglobulins, Antibodies

## Abstract

**Background:** The role of convalescent plasma (COPLA) for the treatment of severely ill Corona Virus Disease-2019 is under investigation. We compared the efficacy and safety of convalescent plasma with fresh frozen plasma (FFP) in severe COVID-19 patients.

**Methods and findings:** This was an open-label, single-centre phase II RCT on 29 patients with severe COVID-19 from India. One group received COPLA with standard medical care (SMC) (n=14), and another group received FFP with SMC (n=15). A total of 29 patients were randomized in the two treatment groups. Eleven out of 14 (78.5%) patients remained free of ventilation at day seven in the intervention arm while the proportion was 14 out of 15 (93.3 %) in the control arm (p= 0.258). The median reductions in RR per min at 48-hours in COPLA-group and FFP group were -6.5 and -3 respectively [p=0.004] and at day seven were -14.5 and -10 respectively (p=0.008). The median improvements in percentage O2 saturation at 48-hours were 6.5 and 2 respectively [p=0.001] and at day seven were 10 and 7.5 respectively (p=0.026). In the COPLA-group, the median improvement in PaO2/FiO2 was significantly superior to FFP at 48-hours [41.94 and 231.15, p=0.009], and also at day-7 [5.55 and 77.01 p<0.001]. We did not find significant differences in hospitalization duration between the groups (0.08).

**Conclusion:** COPLA therapy resulted in rapid improvement in respiratory parameters and shortened time to clinical recovery, although no significant reduction in mortality was observed in this pilot trial. We need larger trials to draw conclusive evidence on the use of Convalescent plasma in COVID-19. This trial is registered with ClinicalTrial.gov (identifier: NCT04346446).

## Introduction

The outbreak of SARS-CoV-2 infection, which had originated in Wuhan, China, has become a pandemic involving more than 35 million population across the world, with almost 1 million deaths and still counting.^1^ The case-fatality rate of COVID-19 (Corona Virus disease-2019) has ranged from 1.2-13%.^1,2^ The current evidence-based strategy relies on providing supportive care in mild to moderate cases and providing mechanical ventilation and extracorporeal membrane oxygenation in severe cases. There is no targeted drug therapy available at present. Some studies have indicated benefits with intravenous remidisvir and a combination of lopinavir and ritonavir in reducing the severity and duration of illness, but not mortality.^3-5^ Till now use of the only dexamethasone has resulted in reduced 28-day mortality among severe and critical COVID-19 patients.^6^ Apart from antiviral drugs, virus-specific neutralizing antibodies, which could accelerate virus clearance and prevent entry into target cells, could serve as a mechanism for the restriction and clearance of the viruses by the host. The plasma of convalescent patients who have recovered from SARS-CoV-2 infection may contain such neutralizing antibodies which may accelerate virus clearance in an infected recipient and be used in the treatment of patients with COVID-19.^7^ The experience of using convalescent plasma is derived from its utility in improving the survival rate of patients with SARS infection wherein the patients who had no response to intravenous corticosteroids showed improvement. Providing passive antibody therapy through convalescent plasma in COVID-19 infection could be one of the approaches towards disease mitigation in the absence of definitive treatment.^5-9^ This approach can be effective in patients before they develop a humoral response to COVID-19.

## Methodology

### Trial design and study setting

It was an open-labeled, phase II; pilot randomized controlled trial conducted to assess efficacy and safety of convalescent plasma at the Institute of Liver and Biliary Sciences (ILBS) and in collaboration with the Department of Internal Medicine, Lok Nayak Hospital, New Delhi, a designated COVID-19 treatment centre. The first Donor was recruited on 20 April 2020 while, the first patient was recruited on 21 April 2020, and final follow up was completed on 30 May 2020. No scientific sample size calculation was done as it was a pilot trial only. A total of 29 patients were included in the study. The trial was approved by the Institutional Ethical Committees and was registered with ClinicalTrial.gov (identifier: NCT04346446). The randomization was done by using block randomization method with varying block sizes by an independent statistician. The allocation concealment was done by using Sequentially Numbered Opaque Sealed Envelopes (SNOSE) method.

Patients were enrolled for convalescent plasma transfusion along with the standard treatment protocol in one arm and fresh frozen plasma [FFP] along with the standard treatment protocol in another arm by random allocation. FFP transfused in the study was collected before the emergence of the virus in our country to avoid any chance of providing COVID-19 convalescent plasma in the SMT group. The primary outcome measure was the proportion of patients remaining free of mechanical ventilation in both groups on day seven. The secondary outcome measures included mortality at day seven and day 28, improvement in PaO2/FiO2, and the SOFA scores reduction at 48 hours and day 7, duration of hospital stay, duration of Intensive Care Unit stay, requirements of vasopressors and days free of dialysis up to 28 days from randomization. In addition to above, clinical assessment of patients was done by assessing reduction in respiratory rate, and improvement in oxygen saturation at 48 hours and seven days and laboratory effects of plasma therapy by improvement in lymphocyte count Ct value at seven days and any adverse transfusion events with plasma transfusion.

We collected 500ml Convalescent plasma (COPLA) from COVID-19 recovered patients after 14 days of complete resolution of symptoms by Plasmapheresis (MCS+, Hemonetics USA) after due consent. Two consecutive test negative results of Real-time reverse transcriptase Polymerase chain reaction (RT-PCR) were done 24 hours apart from combined oral and nasopharyngeal swab for SARS CoV-2 for donation consideration. Final eligibility was ascertained thorough medical history, physical examination and laboratory tests, as per the Drugs and Cosmetics Act, 1940 and further amended on 11.03.2020.^10^

Laboratory testing included serum protein and CBC; transfusion-transmitted infections (hepatitis B virus, hepatitis C virus, HIV, malaria, and syphilis), blood grouping and antibody screening. The collected convalescent plasma was separated into two equal aliquots for divided dosing, labelled as per regulatory requirements and stored at -80°C in a separately designated deep freezer. It was issued to the interventional treatment arm patients under study. FFP, a licensed product collected from whole blood donation from healthy blood donors, was transfused to the control treatment arm.

### IgG antibody against SARS CoV-2 assay

We tested each convalescent plasma unit for the presence of IgG antibodies and neutralizing antibodies to SARS CoV-2. The titres of spike protein S1 RBD IgG antibody was done by ELISA (SARS-CoV-2 Spike S1-RBD IgG Detection Kit, Genscript, USA) representing spike protein antibody, directed against the SARS-CoV-2 RBD (receptor binding domain) proteins. The titre was determined by ELISA using positive, negative controls and sample dilutions (1:80, 1:160, 1:320, 1:640 and 1:1000. All samples were run in duplicate. The ELISA titres were determined by endpoint dilution. Also, the titres in 14 COPLA recipients and 15 FFP recipients were determined after plasma therapy.

### Neutralizing antibodies against SARS-CoV2 assay

The determination of serum neutralization antibodies in donors was done by SARS-CoV-2 Surrogate Virus Neutralization Test (sVNT) Kit (Genscript, USA). The test was used to detect circulating neutralizing antibodies against SARS-CoV-2 that block the interaction between the receptor-binding domains of the viral spike glycoprotein (RBD) with the ACE2 cell surface receptor. The S1 RBD IgG titre of 1:80 or above was preferred. The absorbance of the sample was inversely dependent on the titre of the anti-SARS-CoV2 neutralizing antibodies.

## RT PCR for SARS CoV2

### Sample Collection

Both nasopharyngeal and oropharyngeal swabs were taken and transported in a 3 ml viral transport media, maintaining a proper cold chain to the Virology laboratory. A volume of 200 microlitres (µl) of the sample was further processed for viral nucleic acid extraction by Qiasymphony DSP Virus/ Pathogen mini kit (Qiagen GmbH, Germany) as per the manufacturer’s protocol.

### Performance of RT-PCR

The extracted elutes of RNA was subjected to RT-PCR for the qualitative detection of both E as well as ORF 1ab (RdRP) genes of SARS-CoV-2 virus using a commercial RT PCR kit (nCoV RT–PCR, SD Biosensor, Gyeonggi-do, Republic of Korea) as per the kit literature. The sample was considered positive when fluorescence was seen in both the target genes E as well as RdRP up to a cycle threshold (Ct) value 36. Ct value for each gene was noted separately and keeping in mind that there exists an inverse relationship between Ct value and the amount of viral nucleic acid in the specimen, Ct value was utilized as a marker to monitor the clinical progress of the patients.

### Patient selection

A written informed consent was taken from all the patient at the time of enrolment in the study. The inclusion criteria of the trial were, a SARS-CoV-2 infection (positive by real-time PCR assay) patient, with severe COVID-19 [respiratory rate (RR) ≥30/min, oxygen saturation level less than 93% in resting state, the partial pressure of oxygen (PaO2)/oxygen concentration (FiO2) ≤300 mmHg, lung infiltrates >50% within 24 to 48 hours]. The exclusion criteria were; failure to obtain informed consent, patients less than 18 years or more than 65 years of age, those with co-morbid conditions (cardiopulmonary disease-structural or valvular heart disease, coronary artery disease, COPD, chronic liver disease, chronic kidney disease), patients presenting with multi-organ failure or on mechanical ventilation, pregnant females, individuals with HIV, viral hepatitis, cancer, morbid obesity with a BMI>35 kg/m2, extremely moribund patients with an expected life expectancy of <24 hours, hemodynamic instability requiring vasopressors, previously known history of allergy to plasma, or a PaO2/FiO2 ratio less than 150. The patients selected for inclusion were randomized to receive either transfusion of COPLA or FFP along with the standard treatment protocol within three days of onset of symptoms of severe COVID-19.

### Clinical and Laboratory Monitoring

Demographic details, date of hospital admission, symptoms present at the time of testing, the severity of illness, ongoing treatment protocols, mechanical ventilator support were noted for all patients. Clinical data, including body temperature, PaO2/FiO2, and Sequential Organ Failure Assessment (SOFA) Scores, were collected and analysed. Samples from all potential plasma recipients for baseline and follow-up laboratory monitoring like CBC, serum antibody titres and cytokine levels were obtained. Radiological data of chest x-ray, chest CT were collected, wherever available. Patients were followed-up for any complications like pneumonia, acute respiratory distress syndrome (ARDS), and the development of multi-organ dysfunction syndrome (MODS).

## Treatment protocol and patient monitoring

### Standard of Care

All the patients in the study were initiated on supplemental oxygen at five litter/min with target SpO2 being ≥94%. If saturation remained below 94%, either of high flow Oxygen or NIV (via BiPAP) was given. Medically, all patients received a course of Hydroxychloroquine 400 mg BD on Day1, followed by 200 mg BD for five days along with Oral Azithromycin 500 mg OD for five days. Standard medications for the control of diabetes and hypertension were given when required.

### Management of ARDS

In cases where patients had developed ARDS, were initiated on high flow oxygen (5 - 15 litre/min) and responses were assessed regularly by using a pulse-oximeter and ABG analysis. Still, if the patient remained in hypoxemia, we considered Non-invasive ventilation (NIV) in the form of BiPAP. Despite all this, if no clinical response was observed, elective intubation was done, and patients were placed on mechanical ventilation.

### Transfusion protocol

We transfused ABO blood group compatible 500 ml plasma (either convalescent or FFP) in two divided doses on consecutive days to avoid transfusion-related volume overload. All the patients were regularly monitored for vital signs before, during, and after transfusion to detect any transfusion related adverse events.

### Monitoring of patients

Patients were admitted to the ICU based on the severity of the illness. Daily monitoring and clinical assessment were done by taking vitals (Blood Pressure, Heart Rate, Temperature, Respiratory Rate) after following all IPC protocol. Due to the nature of the COVID-19, monitoring via ABG (Arterial Blood Gases) and pulse-oximetry were done.

After plasma (COPLA or FFP) transfusion, samples for laboratory monitoring on days 0, 3, 7, 14, and day 28 (wherever possible) were collected. Complete Blood Count for analysis of haemoglobin (Hb), Hematocrit (Hct), total leukocyte counts (TLC), and Neutrophils/lymphocytes (N/L) ratio was performed by using electrical impedance based on multi parts cell counter (LH750, Beckman Coulter, Florida, USA). Plasma levels of IL-1β, IL-6, TNF-α, and IL-10 were measured using multiplex procataplex (Thermo Fisher Scientific, Bender MedSystems GmbH, Campus Vienna Biocenter 2, Vienna Austria) using cytokine bead assay following the Complete details provided by manufacturer’s protocol. The Luminex assay was run in xponent3.1TM Rev. 2 (Luminex Corporation, 12212 Technology Boulevard, Austin, Texas, USA) and levels were determined.

### Statistical analysis

Continuous variables were expressed as mean (SD) or median (range) and compared by Student’s t-test or Mann-Whitney U test as appropriate. The categorical data were analyzed using Chi-Square or Fisher’s exact test. To compare the pre and post outcome values, a paired t-test or Wilcoxon signed-rank test was used. To find out the predictor in survival analysis Cox-proportional hazard regression analysis was applied. The actuarial probability of survival was calculated by the Kaplan-Meier graph and compared by the log-rank test. The p-value < 0.05 was considered statistically significant. All statistical tests were performed using SPSS for Windows version 22 (SPSS IBM Corp. Ltd. Armonk, NY)

## Results

A total of 292 patients with COVID-19 were assessed for inclusion in the study, of which 241 were excluded as they had mild to moderate disease and only 51 patients with severe COVID-19 were found eligible. Twenty patients were further excluded due to signs/symptoms of severe COVID-19 for more than three days before randomization (n=13), lack of consent (n=7), or other exclusion criteria. Thirty-one patients underwent randomization out of which 15 were assigned to receive convalescent plasma with standard care and 16 to FFP with standard care. However, in both groups, one patient became RT-PCR negative for SARS-CoV-2 on the day of plasma transfusion. Thus, a total of 14 patients received COPLA and 15 received FFP (Fig. 1).

**Figure 1.**
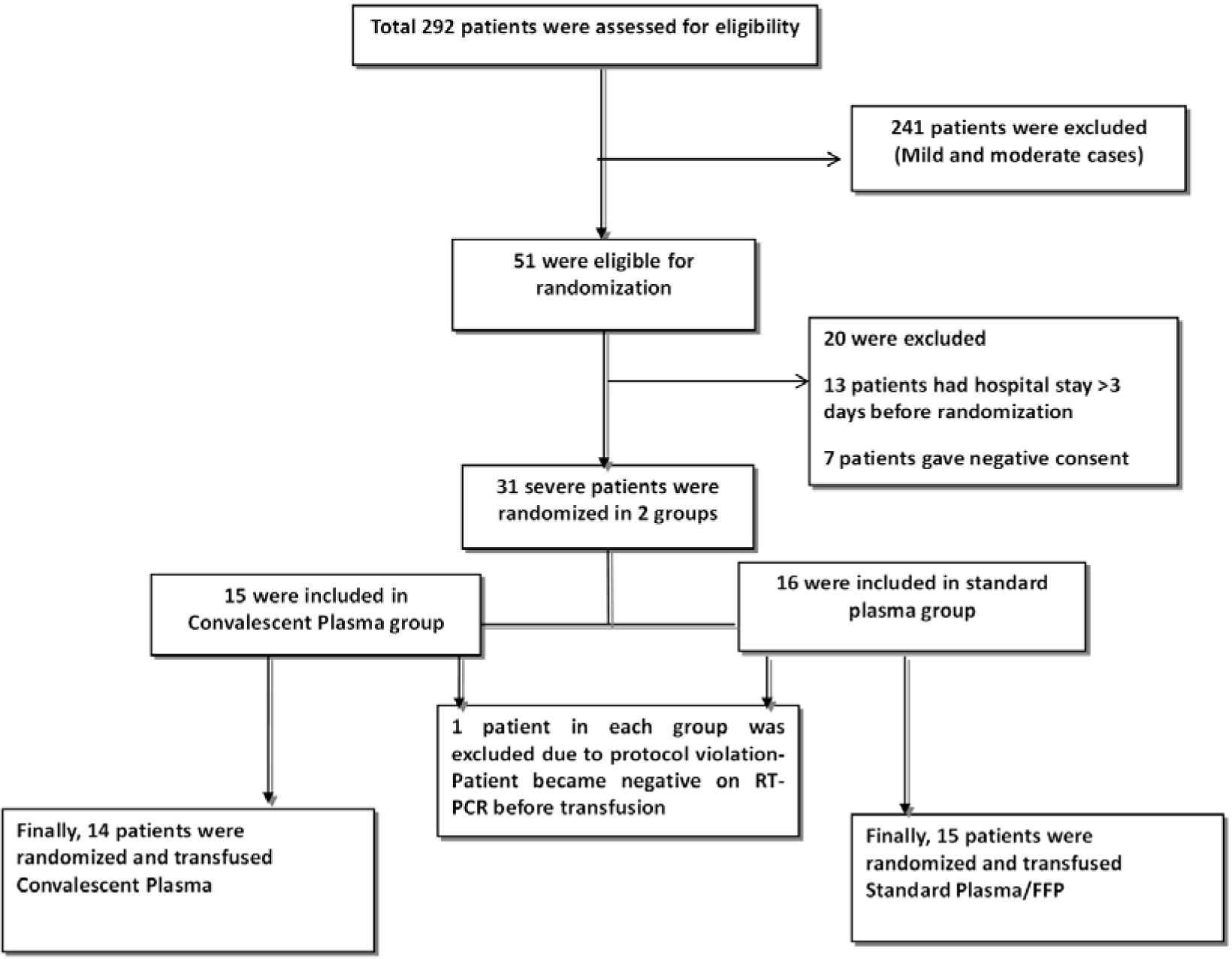
Patient Selection and Randomization.

### Baseline patient profile

The mean age, gender, and body mass index (BMI) were comparable between the groups (Table 1). The patients in both COPLA and FFP groups had severe disease with the high respiratory rate (35.36 ± 2.65 and 34.47 ± 2.47 respectively), and low saturation (% O2 saturation of 85 ± 4.29 and 85.07 ± 3.92 respectively). Baseline PaO2/FiO2 ratios were low (164.92 ± 15.85 and 161.06 ± 11.77 respectively), and X-ray changes were present in 12 (85.7%) and 13 (86.67%) patients respectively in COPLA and FFP groups. The baseline SOFA scores were also comparable.

**Table: 1.**
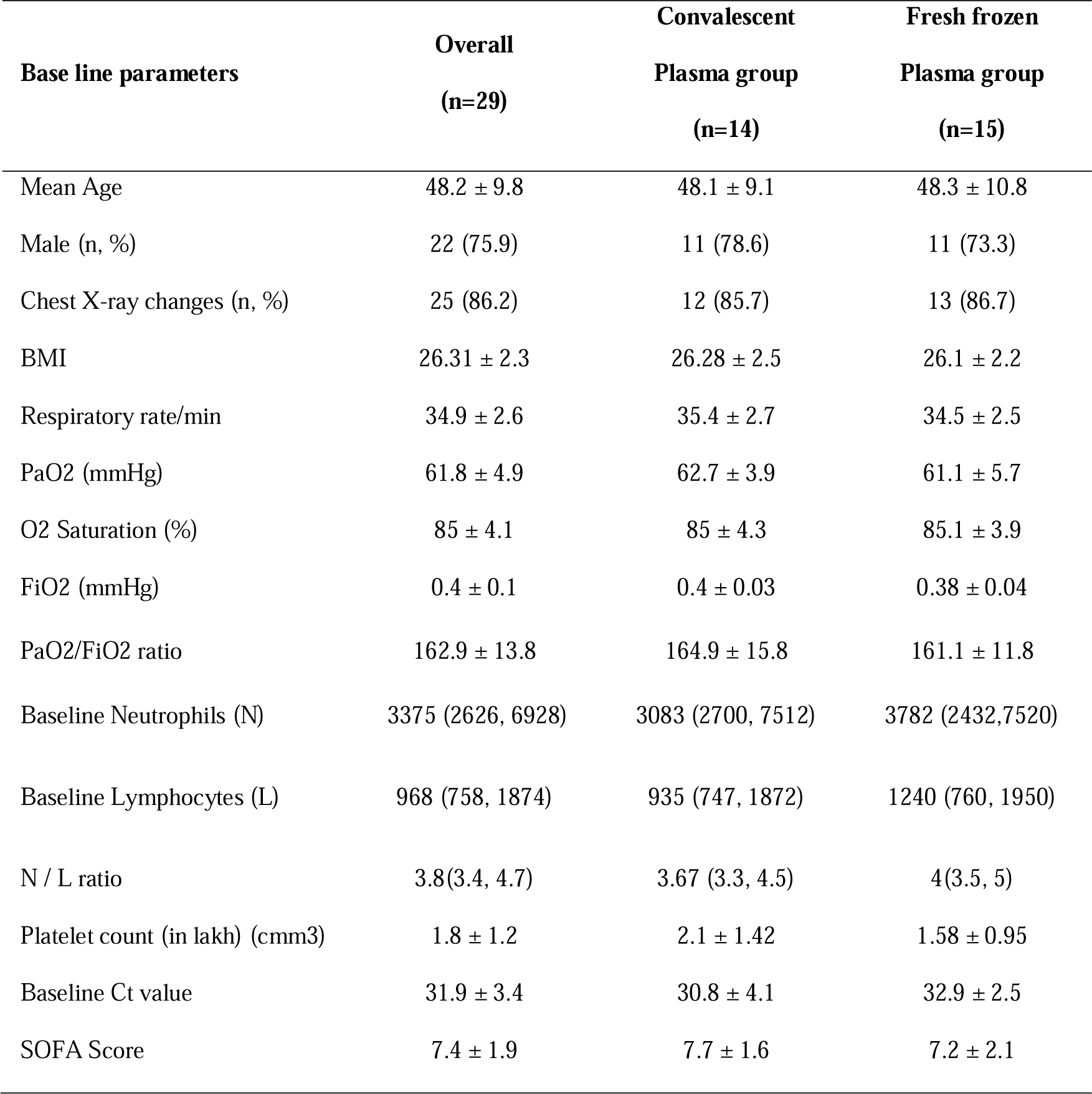
Baseline characteristics of patients under study.

### Convalescent Plasma Donation

Out of 28 COVID-19 positive recovered donor consented for participation, only 14 donors were deemed eligible to donate. The procedural details are described in table 2. None of the donors experienced any adverse donor reaction during the procedure, and no complaint was received on a follow-up after two weeks of donation.

**Table: 2.** Convalescent Plasma Donation Details (n=14)

| S. N. | Parameter | Value |
| --- | --- | --- |
| 1 | Median Age (years) | 33 ( 21, 48 ) |
| 2 | Gender (M /F) | 14 / 0 |
| 3 | Median period between resolution of symptoms and donation (days) | 27 ( 17, 73) |
| 4 | Median Blood Volume Processed (ml) | 1643.5 (1202, 1979) |
| 5 | Median volume of plasma collected | 500 ml (442, 500) |
| 6 | Donor median neutralizing antibody titre | $\geq 80$ (10, $\geq 80$) |
| 7 | Donor median S1 RBD IgG antibody titre | $\geq 640$ (10, $\geq 640$) |
| 8 | Adverse Donor Reactions (ADRs) | None |

## Laboratory parameters

### Baseline laboratory parameters

The baseline N/L ratio in COPLA and FFP groups [7.71 ± 1.59 and 7.20 ± 2.11] and platelet counts [2.05 ± 1.42 and 1.58 ± 0.95] were comparable. In the COPLA group, the mean baseline Ct value was 30.83 ± 4.07 while in the FFP group, it was 32.93 ± 2.46 [p = 0.12] (Table 1).

### Antibody titres

The S1 RBD IgG, antibody titres of the COPLA donors, ranged from 10 to 640 (Table 2). 64.28% (9/14) of donors had IgG titre of 640, 28.57% (4/14) had a titre of 1:80 and 7.14 % (1/14) had a titre of 1:10. The neutralizing antibody titres of >80 were observed in 13 donors, and one donor had titre <10 (Table 2). The S1 RBD IgG antibody titres of the 14 recipients ranged between 10 and 640 a day after the convalescent plasma transfusion and 0 to 640 after FFP transfusion. There was a significant time-dependent increase in the IgG antibody titres in 85.71% (12 out of 14) of the convalescent plasma recipients as compared to 20% (3 out of 15) FFP recipients (p= 0.001) (Table 4). On intragroup analysis, we found that in COPLA group S1 RBD IgG antibody titres were changed from median titres of 80 to 640 (400% change, p= 0.002) while no difference in median titre of 80 was there in FFP group (p= 0.083).

**Table: 4.** Changes in clinical and laboratory outcome parameters.

| Variable | Convalescent Plasma (n=14) | Normal Plasma (n=15) | Effect Size (95% CI) | P value |
| --- | --- | --- | --- | --- |
| Median Reduction in Respiratory Rate/min at 48 hours | -6.5 (-10.3, -5) | -3 (-5,-1) | -4.2 (-7.8,-0.6) | 0.004 |
| Median Reduction in Respiratory Rate/min at 7 days | -14.5 (-18.7, -13) | -10 (-14,-9) | -3.4 (-6.1,-0.7) | 0.008 |
| Median Improvement in O2 Saturation at 48 hour | 6.5 (5, 7.3) | 2 (1, 2) | 4.6 (2.9, 6.18) | <0.001 |
| Median Improvement in O2 Saturation at 7 days | 10 (8.2, 11) | 7.5 (4.7, 9.3) | 2.8 (0.2, 5.5) | 0.026 |
| Mechanical Ventilation required within 7 days (n) % | 3 (21.4) | 1 (6.7) | 3.2 (0.4, 27.4)* | 0.26 |
| Median reduction in SOFA Score 48 hours | -2 (-2.3, -1) | -1 (-1, 0.0) | -0.54 (-1.9, 0.9) | 0.01 |
| Median Reduction in SOFA Score 7 days | -5 (-6.5,-4.0) | -3 (-5.3, -2.7) | -0.8 (-2.8, 1.6) | 0.04 |
| Median improvement in PaO2/FiO2 at 48 hour | 41.9 (1.3, 55.6) | 5.55 (-9.3, 11.1) | 32.5 (8.3, 56.7) | 0.009 |
| Median improvement in PaO2/FiO2 at 7 days | 231.2 (183.4, 245.2) | 77.0 (56.9, 96.2) | 30.9 (51.4, 183.9) | <0.001 |
| Median ICU stay | 5 (4, 5.7) | 5 (4,7) | -0.13 (-2.9, 2.7) | 0.72 |
| Mean duration of Hospital stay (days) | 12.1 ± 4.1 | 16.1 ± 5.6 | -3.9 (-7.7, -0.2) | 0.08 |
| Vasopressors requirement till 28 days (n) % | 3 (21.4%) | 1 (6.7%) | 3.8 (0.3, 41.9)* | 0.33 |
| Increase in S1 RBD IgG antibody titre post transfusion | 12 (85.7%) | 3 (20%) | 4.3 (1.5, 12.1)* | <0.001 |
| Median Improvement in Lymphocyte count at 7 days | 896.5 (524.5, 1351.5) | 634.0 (218.0, 931.5) | 258.1 (-158.2, 674.3) | 0.15 |
| Transfusion reactions (n) | 1 (7.1%) | 1 (6.7%) | 1.1 (0.1, 15.5)* | 1 |
| Median Improvement in Ct value at 7 days | 7.7 (3.4, 9.2) | 5.2 (3.3, 6.3) | 2.2 (-0.6, 5.1) | 0.11 |
| Mortality till 7 days (n) % | 2 (14.3%) | 1 (6.7%) | 2.1 (0.2, 21.1)* | 0.59 |
| Mortality till 28 days (n) % | 3 (21.4%) | 1 (6.7%) | 3.2 (0.4, 27.4)* | 0.33 |

Risk Ratio (95% CI)

### Ct Value

The change in the Ct value represents a reduction in the viral load. It was measured with respect to the rise in the antibody titres in the recipients, and it was observed that the median increment in the Ct values at day seven was 7.7 (3.4, 9.2)and 5.2 (3.3, 6.3) in the COPLA group and FFP group respectively. There was a higher reduction in the viral load in the COPLA group, but possibly due to the small sample size, the difference was not significant (p=0.11) (figure 2, table 4).

**Figure 2.**
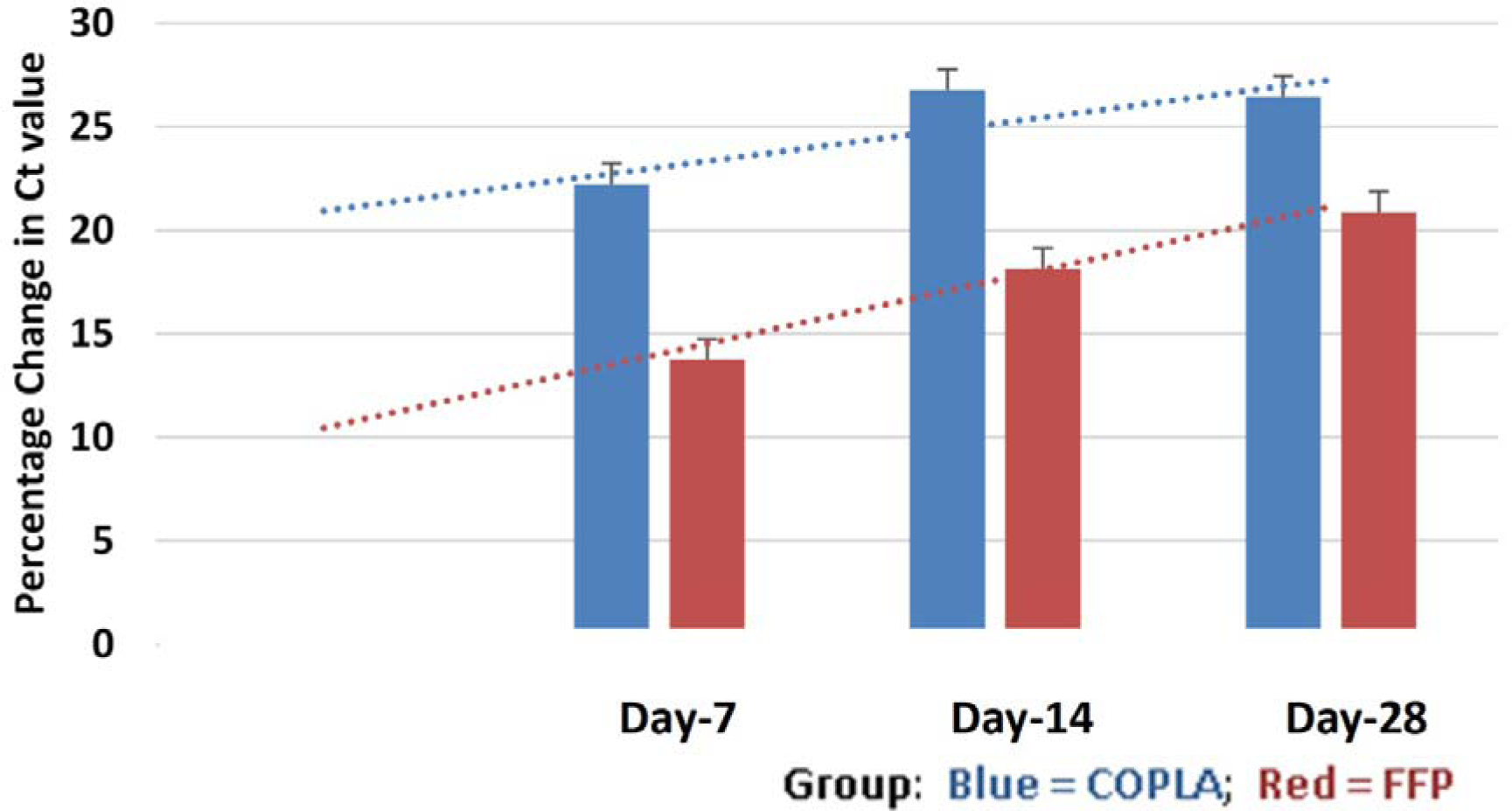
Reduction in viral load after transfusion (Ct value)

### Cytokine levels

The baseline cytokine levels were comparable in both the study arms. On convalescent plasma transfusion, median post-transfusion IL-6, IL-10, and TNF-α levels were reduced while IL-1β level was increased at day 7. In the FFP group, the median post-transfusion levels of IL-1β and TNF-α were reduced while IL-6 and IL-10 showed an increase. These differences did not attain statistical significance post-transfusion, as shown in table 3.

**Table: 3.** Median baseline and post transfusion cytokine level.

| Cytokines | COPLA-Baseline | COPLA- Post transfusion | p-value | FFP-Baseline | FFP-Post transfusion | p-value |
| --- | --- | --- | --- | --- | --- | --- |
| IL-1β | 4.2 (2.1, 12.3) | 9.9 (3.3, 23.1) | 0.75 | 3.3 (1.6, 11.1) | 2.8 (1.6, 7.3) | 0.31 |
| IL-6 | 52 (17.1, 219.4) | 31.8 (26.9, 104.9) | 1 | 45.5 (16.4, 62.2) | 49.8 (21.8, 341.2) | 0.31 |
| IL-10 | 4.5 (1.3, 7.1) | 3.3 (1.5, 4.3) | 0.86 | 3.1 (1.9, 5.3) | 4.7 (3.4, 5.3) | 0.17 |
| TNF-α | 7.0 (7.0, 9.7) | 6.0 (5, 7) | 0.18 | 7.0 (7, 7) | 6 (5, 7) | 0.28 |

### Clinical evaluation

The primary outcome of clinical efficacy was objectively analyzed in both the treatment groups. Three (21.4%) patients in the COPLA group and 1 (6.7%) in the FFP group needed mechanical ventilation within seven days of transfusion; the proportion of patients free of ventilation at day seven were 11 out of 14 in the COPLA arm and 14 out of 15 in the control arm, this difference was not statistically significant (p= 0.258) [Table 4]. The secondary outcome measures showed significant benefits of convalescent plasma. In the COPLA group compared to the FFP group, the median reductions in respiratory rate per min at 48 hours were -6.5 (−10.3, -5) and -3 (−5,-1) respectively [p=0.004] and at day 7 were -14.5 (−18.75, -13) and -10 (−14, -9) respectively [p=0.008]. In the COPLA group, the median improvement in % O2 saturation at 48 hours and day 7 was 6.5 (5, 7.25) and 10 (8.2, 11) respectively. In the FFP group, the median improvement in % O2 saturation at 48 hours and day 7 was 2 (1, 2) and 7.5 (4.75, 9.25) respectively (p = < 0.001, and p = 0.02). In the COPLA group, the median reduction in SOFA score at 48 hours and day 7 was -2 (−2.25,-1) and -5 (−6.5,-4.0) respectively while it was -1 (−1, 0.0) and -3 (−5.25, -2.75) respectively in the FFP arm (p=0.016, and p=0.047). In the COPLA arm, median improvement in PaO2/FiO2 at 48 hour and at 7 days was 41.94 (1.25, 55.58) and 231.15 (183.37, 245.2) respectively compared to 5.55 (−9.32, 11.11) and 77.01 (56.93, 96.20) respectively in the FFP arm (p=0.009, and p = < 0.001). We found an early and statistically significant reduction in respiratory rate/min, improvement in O2 saturations, reductions in SOFA scores and PaO2/FiO2 at 48 hours and seven days in the COPLA group as compared to the FFP group. The lymphocyte count was higher y t in the COPLA group on day 7. Requirements of vasopressors were required overall in 4 patients (3 in the COPLA group and 1 in the FFP group) until 28 days, but it not significant statistically (p=0.33) [Table 4].

In COPLA group median ICU stay, and mean hospital stay was 5 (4, 5.7) and 12.1 ± 4.27 days respectively, while in the FFP group they were 5 (4, 7) and 16.1 ± 5.6 days respectively. These differences were not significant statistically (p= 0.72, and p= 0.08).

### Mortality

A total of 4(13.79%) out of 29 patients, succumbed to COVID-19 during the study period of 28 days out of which three were in COPLA group, and one was in FFP group. One patient in the COPLA group had a severe acute respiratory illness with oliguria, uremic encephalopathy, and AKI at presentation. Ideally, due to multi-organ failure, he should not have been included, but since some of the laboratory reports came after randomization and transfusion. This patient was put on haemodialysis. He became virus-negative by RT–PCR 6 days after the plasma therapy but succumbed on day 15 due to progressive multi-organ failure. Two other patients who died in the COPLA group showed a continuous fall in O2 saturation from the time of randomization to transfusion, and thereafter, thus, further transfusion was withheld, and the patients succumbed within 24 hours. One patient in the FFP group, one 60 years male, showing SARI was admitted with poor O2 saturation and put on ventilator support. He received FFP on day three and day four but died due to cardiac arrest on day 5. Both the treatment arms were compared for survival analysis using Cox-proportional hazard regression. In COPLA group three events of mortality were observed while in the FFP group, one event was observed, the difference was statistically not significant (HR, 4.23 [95% CI, 0.43-41.6]; P = 0.22).

### Safety profile

One patient in each of the arms, showed signs of mild urticaria during plasma transfusion, which was comfortably managed by treating physician. No significant untoward side effects of convalescent plasma or FFP transfusion were observed in the study (p= 1), as shown in Table 4.

## Discussion

We observed the safety of both types of plasma (convalescent plasma and FFP) and found that convalescent plasma is as safe as FFP. Hence, convalescent plasma can be transfused to COVID-19 patients without any added potential risk similar to the findings reported by Joyner et al. on 5000 patients who were transfused COPLA and found it to be a safe treatment modality in COVID-19 patients.^11^ The preliminary results in the present pilot trial are encouraging and seem promising as clinical recovery by an early and significant improvement in O2 saturation; reduction in respiratory rate and reduction in SOFA scoring was observed. We found shorter ICU, and hospital stays using convalescent plasma, but these findings were not significant. Still, there is no established therapy at present for severe COVID-19. There are equivocal reports of plasma therapy compared to no therapy for patients with severe Covid-19. The results of our trial show that convalescent plasma significantly reduced the respiratory rate, improved O2 saturation, and improved the PaO2/FiO2 ratio compared with fresh frozen plasma. There was no difference; however, in the number of patients requiring ventilation or on mortality.

In this novel study, the purpose of using FFP in the control arm was to supplement and balance the beneficial effects of plasma, whether FFP or COPLA, on coagulation abnormalities developing in severe COVID-19 patients and study the added benefits specific to SARS-CoV-2 antibodies present in COPLA.

Use of Convalescent plasma therapy in severe COVID-19 infection could be one of the approaches towards disease mitigation in the absence of definitive treatment. There were clear advantages of giving COPLA over FFP as evident from the results of our study. The convalescent plasma therapy improved respiratory clinical parameters, but did not impact the need for ventilation and also it did not improve survival. Thus its role in severe patients who require ventilator support and who show rapid deterioration is not clear. Similar findings were observed in a recently conducted trial in the China by Ling et al. where they didn’t find a statistically significant improvement in severe and critically ill COVID-19 patients in time to clinical improvement within 28 days.^12^ Contrary to this, convalescent plasma therapy has shown improved survival in anecdotal and small case series; in five critically ill patients by Shen et al., in 10 critically ill patients by Duan et al. and in 4 critically ill patients by Zhang et al. respectively.^5,8,9^

There was a significant time-dependent increase in the S1 RBD IgG antibody titres in patients who received COPLA as compared to FFP this was similar to the study done by Shen C et al. where they found an increase in S1 RBD IgG antibody titres after COPLA transfusion.^5^ In this pilot trial, we found an early increase in the Ct values in the COPLA group, demonstrating a speedy reduction in the viral load. We found an early reduction in viral load in the convalescent plasma group as compared to the FFP group. RT-PCR results showed no detectable SARS CoV-2 RNA target in the nasopharyngeal swab within 14 days in the COPLA group as compared to the FFP group where it took more than 14 days to become negative. It is a critical laboratory marker for assessing the effectiveness of therapy, which corroborates with all the previous studies done on it.^13, 14^

Further, the appearance of the anti-inflammatory marker IL-10 and reduction in levels of pro-inflammatory markers (IL-1, IL-6, and TNF-TNF-α), has been well correlated with the disappearance of clinical symptoms.^15^ We found a decrease in IL-6 and TNF-α level post-transfusion, showing it can limit immune-mediated damage and associated complications and early sign of recovery in COVID-19 patients in the convalescent plasma group.^16^ In the FFP group, we found an isolated decrease in the TNF-α level on transfusion while an increase in the IL-6 showing ongoing active corona infection with the beneficial effect of FFP on endothelium lining and coagulation system, as shown in a study done by Straat et al.^17^

Our study had certain limitations. We had fewer numbers of participants in this pilot trial thus we could not draw clear cut and robust conclusions. Secondly, one patient in the convalescent plasma group was already in renal failure. Third, all the patients were given Oseltamivir anti-viral and hydroxychloroquine despite the uncertainty of the efficacy as part of their standard care. Lastly, the dynamic changes in cytokines level during treatment were not investigated in an absolute controlled way. However, the preliminary results in the present pilot trial are encouraging and seem promising as early clinical recovery and shorter ICU, and hospital stay was observed in the convalescent plasma group. We need to conduct larger RCTs to draw conclusive evidence before advocating this mode of therapy.

## Data Availability

Data is available with Project Investigators at Institute of Liver and Biliary Sciences, and Maulana Azad Medical College and will be available with permission of IRB of above institute

## Acknowledgements

We thank Mr Pankaj Jain for his technical support in the trial.

## Supporting Information files

1. Statistical analysis plan
2. Protocol
3. Addendum in protocol
4. Consort Checklist

